# Associations of Serum GDF-15 Levels with Physical Performance, Mobility Disability, Cognition, Cardiovascular Disease, and Mortality in Older Adults

**DOI:** 10.1101/2024.08.07.24311629

**Authors:** Katey Webber, Sheena Patel, Jorge R. Kizer, Richard Eastell, Bruce M. Psaty, Anne B. Newman, Steven R. Cummings

## Abstract

**Background:** Growth differentiation factor 15 (GDF-15) is a member of the TGFβ superfamily secreted by many cell types and found at higher blood concentrations as chronological age increases (1). Given the emergence of GDF-15 as a key protein associated with aging, it is important to understand the multitude of conditions with which circulating GDF-15 is associated.

**Methods:** We pooled data from 1,174 randomly selected Health ABC Study (Health ABC) participants and 1,503 Cardiovascular Health Study (CHS) participants to evaluate the risk of various conditions and age-related outcomes across levels of GDF-15. The primary outcomes were (1) risk of mobility disability and falls; (2) impaired cognitive function; (3) and increased risk of cardiovascular disease and total mortality.

**Results:** The pooled study cohort had a mean age of 75.4 +/−4.4 years. Using a Bonferroni-corrected threshold, our analyses show that high levels of GDF-15 were associated with a higher risk of severe mobility disability (HR: 2.13 [1.64, 2.77]), coronary heart disease (HR: 1.47 [1.17, 1.83]), atherosclerotic cardiovascular disease (HR: 1.56 [1.22, 1.98]), heart failure (HR: 2.09 [1.66, 2.64]), and mortality (HR: 1.81 [1.53, 2.15]) when comparing the highest and lowest quartiles. For CHS participants, analysis of extreme quartiles in fully adjusted models revealed a 3.5-fold higher risk of dementia (HR: 3.50 [1.97, 6.22]).

**Conclusions:** GDF-15 is associated with several age-related outcomes and diseases, including mobility disability, impaired physical and cognitive performance, dementia, cardiovascular disease, and mortality. Each of these findings demonstrates the importance of GDF-15 as a potential biomarker for many aging-related conditions.

## INTRODUCTION

Aging biomarkers have come to the forefront in the study of aging-associated diseases and disabilities common in older adults. Unlike chronological age, specific aging biomarkers target the changes on a cellular level and offer potential opportunities to reduce disease and disability among older adults by intervening on biological pathways reflected by specific markers. Growth differentiation factor 15 (GDF-15) has the potential to be an important biomarker as it is strongly associated with chronological age (1) and several aging-related outcomes.

A member of the TGFβ super family, GDF-15 is secreted by many cell types. It is found at higher circulating concentrations as chronological age increases, particularly after age 65, and is upregulated by stress (1). Two studies measuring protein concentrations have identified GDF-15 as a protein strongly associated with conditions that are believed to induce cell senescence (2,3). These studies suggest that GDF-15 may be a senescence-associated secretory phenotype (SASP) factor, reflecting the quantity of senescent cells. Yet, the precise stimuli that increase GDF-15 and the mechanisms for its diverse effects are not known.

Despite the precise mechanisms being unknown, it is important to explore the associations with certain age-related outcomes and other markers of health status. Variation in the circulating level of GDF-15 has been associated with many aging-related diseases, including cardiovascular disease and mortality (4). Previous studies have also shown associations between higher GDF-15 levels and poorer performance on tests of physical performance, such as walking speed (5).

Given the various associations with aging-related diseases and mortality, we postulated that higher levels of GDF-15 would be a marker for many conditions that increase with aging, specifically increased risk of falls, increased risk of mobility disability, slower walking speed, impaired cognitive function, increased risk of cardiovascular disease along with increased total mortality. To date, no study has broadly explored in large cohorts of older adults the associations of levels of GDF-15 with a broad range of conditions that change with aging. We tested these associations in cohort studies of aging including the Health ABC Study (HABC) and the Cardiovascular Health Study (CHS) study. We hypothesized that within our cohort we would replicate those known longitudinal associations between GDF-15 and dementia, cardiovascular disease, and total mortality as well as known crossectional associations with cognitive and physical performance. In addition to these known associations, we also postulated that GDF-15 is crosssectionally associated with higher odds of falls and longitudinally associated with higher risk of persistent and severe mobility disability, conditions that have not been evaluated previously.

## METHODS

### Study Populations

These analyses use serum samples and data from community dwelling older adults in the Health, Aging and Body Composition Study (Health ABC) and Cardiovascular Health Study (CHS). Prior publications have described in detail the design, methods, and procedures of CHS (6) and Health ABC (7). To summarize, CHS is a cohort study of community-dwelling adults age 65 years and older recruited from four field sites (Forsyth County, NC; Sacramento County, CA; Washington County, MD; and Pittsburgh, PA). The CHS total sample included an initial cohort of 5201 participants enrolled in the 1989–90 examination, and a second cohort of 687 primarily African-American participants enrolled in the 1992–93 examination. Examinations included medical interviews, physical assessments, biospecimen collection for laboratory testing and long-term storage, and further diagnostic testing following standardized protocols. Semi-annual contacts with participants involved annual exams alternating with telephone calls between 1989-90 and 1998-99, with semi-annual calls continued thereafter. This study uses data and serum samples from the 1994-95 CHS visit (9).

Health ABC is a prospective cohort study of community-dwelling older adults aged 70 to 79 years. From a list of Medicare beneficiaries residing in areas surrounding Pittsburgh, PA and Memphis, TN, 3,075 participants were recruited for a baseline examination in 1997-98. These participants were contacted by telephone every 6 months and attended clinical visits annually or biannually. Visits involved standardized medical questionnaires, physical examination, blood and urine collection, and laboratory and diagnostic testing. During follow-up visits, health status was assessed and data about interim hospitalizations or major outpatient procedures were collected and centrally adjudicated. The present study uses data and serum samples from the 1997-98 visit (9). All participants in CHS and Health ABC provided written informed consent and IRB approval was obtained for each clinical site and coordinating center.

This present study pooled 2,677 potential participants from CHS and Health ABC cohorts. From a total of 4,842 with available samples at the 1994–95 exam, 1,503 CHS participants were randomly selected for inclusion in the present analysis. From a total of 3,075 participants with available samples at Health ABC study baseline, 1,174 were randomly selected.

### Covariates

#### Covariates in Cross-sectional Models

In both cohorts, age, sex, race, and current smoking status were self-reported. Race was dichotomized as white race or not white race. Current smoking status was self-reported as current smoker, past smoker, or never smoker and was dichotomized for this analysis to current smoker or not current smoker. Body mass index was calculated as weight (kg)/height^2^ (m^2^). Heavy alcohol use was defined as greater than 7 drinks per week.

#### Covariates in Longitudinal Models

All longitudinal models include the above covariates as well as the following covariates. Diabetes was defined as fasting glucose >126 mg/dl, non-fasting glucose >200 mg/dl or use of hypoglycemic medication. Systolic blood pressure was measured in the right arm in seated participants after a five-minute rest using a standard sphygmomanometer. Mean forced expiratory volume in 1 second (FEV1) was measured with a water-sealed, Collins Survey II spirometer (WE Collins, Braintree, MA). Fasting triglycerides, HDL, and LDL analyses were performed on an Olympus Demand system (Olympus Corp., Lake Success, NY). eGFR was calculated from serum concentrations of creatinine and cystatin C measured at baseline using the 2012 Chronic Kidney Disease Epidemiology Collaboration equation (10). In both cohorts, medical history of claudication, CHD, stroke and HF was self-reported (11,12). In CHS, this was validated against medical records or questionnaire responses from personal physicians at baseline, with standard adjudication applied for subsequent events preceding the 1994-95 visit (13). Atrial fibrillation was identified by 12-lead ECG or ICD-9 diagnostic code.

### GDF-15 Assays

Blood samples from the two cohorts’ repositories were stored at −70-80° C. GDF-15 was measured in serum by ELISA (R&D Systems; cat number DGD150). The reportable range was 93.7 pg/ml to 6,000 pg/ml. The analytical CV for serum was 1.6%. Because GDF-15 was not normally distributed, results are presented by quartile of GDF-15.

### Longitudinal Outcomes

#### Cardiovascular outcomes

Three cardiovascular outcomes were investigated longitudinally: coronary heart disease (CHD), and atherosclerotic cardiovascular disease (ASCVD), and heart failure (HF). CHD was defined as myocardial infarction, angina, coronary revascularization (angioplasty or bypass surgery), and/or CHD death. ASCVD comprised non-fatal and fatal MI and non-fatal and fatal stroke. HF and HF subtypes, HF with preserved ejection fraction (HFpEF) and HF with reduced ejection fraction (HFrEF) were examined secondarily. In CHS, adjudication of cardiovascular disease was conducted centrally by an events committee based on review of medical records, as previously described (8, 14, 15) and follow-up duration was through 2015 for CHS and 2012 for Health ABC. Event adjudication in Health ABC was modelled on the approach used in CHS, but non-fatal events were adjudicated locally while deaths were centrally by a committee of geriatricians and internists.

#### Mortality

In CHS, deaths were identified by the field centers during surveillance calls, by proxy report, by review of local obituaries and by the Coordinating Center using files from the Center for Medicare and Medicaid Services. The events committee adjudicated cause of death using medical records, informant interviews, and death certificates. In Health ABC, deaths were identified by an adjudication committee reviewing annual in-person examinations and telephone interviews, medical records, obituaries, death certificates, and files from the Centers for Medicare and Medicaid Services. We evaluated total mortality longitudinally.

#### Dementia

Dementia was evaluated differently by each cohort. In CHS, 3608 partcipants with a brain MRI in 1992-93 were classified as high risk or lower risk of dementia in 1998-99. All high risk and a sample of lower risk individuals had a neuropsychological battery, neurological exam, and a neuropsychiatric inventory. High risk of dementia was defined as a 3MSE score <80, a decline of 5 or more points in MMSE, a TIC score of <28 or and IQ code >3.6, a history of stroke, residence in nursing home, belonging to a minority group or having dementia code in a hospital record. Dementia status was adjudicated by a committee of physicians using standard criteria based on the baseline brain MRI, all cognitive testing, medical records and medications over time as well as the neuropsychological, psychiatric and neurological exams (16). In Health ABC, dementia was determined using an algorithm that considered if participants were prescribed dementia medications (galantamine, rivastigmine, memantine, donepezil, and tacrine), had hospital records with dementia as a discharge diagnosis, or had a decline in 3MSE score of more than 1.5 SD from baseline. Due to the differences in determining dementia status, dementia outcomes are presented separately for each cohort. Risk of dementia was evaluated longitudinally, excluding cases of prevalent dementia.

#### Mobility Disability

In Health ABC, incident persistent mobility disability was defined as participants reporting any difficulty walking ¼ mile or climbing 10 steps on at least 2 consecutive 6-monthly contacts.

Incident severe persistent mobility disability was defined as participants indicating an inability or severe difficulty in either walking ¼ mile or climbing 10 steps during the same time frame. These were assessed longitudinally and those who had previously met criteria were not eligible for enrollment in the study.

### Cross-sectional Outcomes

#### Falls

In Health ABC, participants were asked to self-report falling. For analysis purposes, these were categorized into falling two or more times and assessed crosssectionally. History of falls was included as a covariate in the model.

#### Physical performance

In both CHS and Health ABC, usual walking pace over 6 meters (m/s) was assessed. Maximum grip strength in both hands for 2 trials was measured by Jamar dynamometers. CHS and Health ABC each measured endurance differently. Health ABC measured endurance by the time to complete the 400m walk instructing participants to walk “as quickly as possible.” CHS measured endurance using the six-minute walk test measuring the distance covered in six minutes. Health ABC used KinCom devices to measure the isokinetic knee extensor strength at 60 degrees.

In Health ABC, participants were queried every 6 months about ability and difficulty in walking ¼ mile or climbing 10 steps without resting.

#### Cognitive performance

In both cohorts, cognitive function was regularly assessed with the Teng Modified Mini-Mental Status Examination (3MS) and Digit Symbol Substitution Test (DSST) at baseline and annually or semi-annually.

#### Other measures

Visual contrast sensitivity was measured using the Pelli-Robson Contrast Sensitivity Chart and associations were evaluated crossectionally.

### Statistical Analyses

Analyses combined individual-level data across CHS and Health ABC studies. In certain cases, only Health ABC data or only CHS data were available for the outcome of interest. Continuous variables were described by means and standard deviations, and categorical variables by counts and percentages, stratified by cohort. We divided participants into quartiles by serum levels of GDF-15. We compared baseline characteristics for quartiles of levels using *χ*^2^ tests for categorical characteristics and analysis of variance for continuous characteristics. Multivariable Cox proportional hazards models were used to estimate hazard ratios by increase in GDF-15 quartile. For all cross-sectional outcomes, models were adjusted for age, cohort, sex, race, BMI, smoking status, and heavy drinking. Serial models are included in eTable 1 and eTable 2. For longitudinal mortality and cardiovascular outcomes, a fourth model was included which was additionally adjusted for systolic blood pressure, hypertensive medications, diabetes, stroke, claudication, and atrial fibrillation. In addition, triglycerides, HDL, and LDL were added for CHD and mortality; prevalent CHD was added for HF outcomes and mortality. Additionally, HF was added for the mortality outcome. Proportional hazards assumptions and the absence of co-linearity were verified for all models. All analyses were performed with SAS 9.4 (SAS Institute Inc., Cary, NC). All reported P-values are two-sided. We implemented Bonferroni correction for all primary outcomes to account for multiple testing, determining significance at p<0.05/9=0.006 for cardiovascular outcomes, mortality, dementia, and mobility disability.

## RESULTS

### Baseline Characteristics

Baseline characteristics of the two study cohorts are described in Table 1. At the time of blood draw, the average age in the Health ABC study was 73.5 and in the CHS study was 76.9 years. The two cohorts differed in proportions of men and women and BMI (Table 1). The pooled cohort was 45.3% male, 80.9% white, with a mean age of 75.4 years. Table 2 presents baseline characteristics by quartile of GDF-15. More men fell into the higher quartile of GDF-15 levels compared to women. Smoking status differed across levels of GDF-15. There was no association with race or BMI.

**Table 1.** Characteristics of Participants by Cohort.

|  | **Health ABC**<br>(N= 1174) | **CHS**<br>(N= 1503) | **OVERALL**<br>(N= 2677) |
| --- | --- | --- | --- |
| Age | 73.5 +/- 2.9 | 76.9 +/- 4.8 | 75.4 +/- 4.4 |
| Male | 596 (50.8) | 616 (41) | 1212 (45.3) |
| White | 727 (61.9) | 1439 (95.7) | 2166 (80.9) |
| BMI, kg/m<sup>2</sup> | 27.2 +/- 4.5 | 26.4 +/- 4.2 | 26.7 +/- 4.4 |
| Current smoker | 112 (9.5) | 108 (7.3) | 220 (8.3) |
| Heavy drinker | 41 (3.5) | 145 (9.6) | 186 (7) |
| Good/excellent health | 1005 (85.7) | 1210 (80.6) | 2215 (82.8) |
| Avg sitting systolic BP, mm Hg | 135.2 +/- 19.5 | 132.6 +/- 20.1 | 133.7 +/- 19.9 |
| EGFR | 73.1 +/- 18.2 | 66.5 +/- 16.8 | 69.4 +/- 17.7 |
| FEV1 | 2.2 +/- 0.6 | 2.0 +/- 0.6 | 2.1 +/- 0.7 |
| LDL (mg/dL) | 122.1 +/- 34.7 | 128.6 +/- 32.7 | 125.7 +/- 33.7 |
| HDL (mg/dl) | 53.4 +/- 17.0 | 53.1 +/- 14.0 | 53.3 +/- 15.4 |
| Triglyceride (mg/dl) | 139.5 +/- 80.4 | 146.2 +/- 85.0 | 143.2 +/- 83.1 |
| Teng 3MS score | 90.8 +/- 7.7 | 91.5 +/- 8.5 | 91.2 +/- 8.2 |
| Digit symbol substitution | 36.4 +/- 14.1 | 41.3 +/- 13.0 | 39.2 +/- 13.7 |
| 6m gait speed, m/s | 1.2 +/- 0.2 | 0.9 +/- 0.2 | 1.0 +/- 0.3 |
| Max grip strength, kg | 33.4 +/- 10.7 | 30.1 +/- 10.1 | 31.5 +/- 10.5 |
| 400m walk time | 318.0 +/- 72.5 | - | 318.0 +/- 72.5 |
| Six min walk distance | - | 1143.9 +/- 284.1 | 1143.9 +/- 284.1 |
| Leg power | 94.3 +/- 33.8 | - | 94.3 +/- 33.8 |
| Log contrast sensitivity | 1.6 +/- 0.2 | - | 1.6 +/- 0.2 |
| Diabetes | 205 (17.5) | 227 (16.2) | 432 (16.8) |
| Anti-hypertensive medication | 631 (54) | 761 (50.7) | 1392 (52.1) |
| History of Coronary Heart Disease | 231 (19.7) | 360 (24) | 591 (22.1) |
| History of Atrial Fibrillation | 40 (3.4) | 143 (9.5) | 183 (6.8) |
| Two or more falls | 74 (7.1) | 139 (9.7) | 213 (8.6) |
| GDF-15, pg/mL | 1128.7 +/- 496.0 | 1282.9 +/- 629.3 | 1215.2 +/- 579.6 |

Note. Means +/- SD and n (%) presented.

**Table 2.**
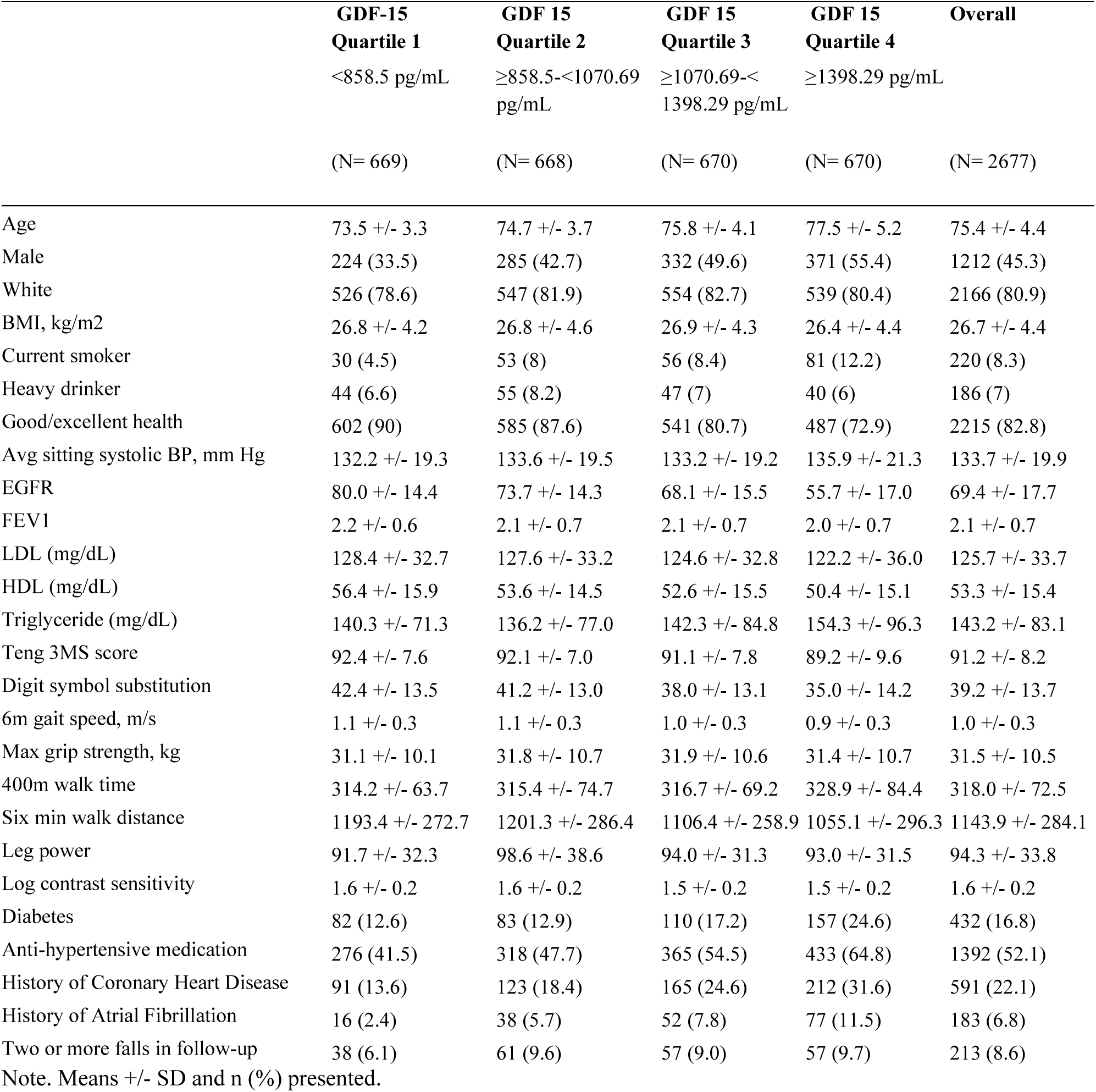
Characteristics of Participants by Quartiles of GDF-15.

|  | **GDF-15**<br>**Quartile 1**<br><858.5 pg/mL<br>(N= 669) | **GDF 15**<br>**Quartile 2**<br>≥858.5-<1070.69<br>pg/mL<br>(N= 668) | **GDF 15**<br>**Quartile 3**<br>≥1070.69-<<br>1398.29 pg/mL<br>(N= 670) | **GDF 15**<br>**Quartile 4**<br>≥1398.29 pg/mL<br>(N= 670) | **Overall**<br>(N= 2677) |
| --- | --- | --- | --- | --- | --- |
| Age | 73.5 +/- 3.3 | 74.7 +/- 3.7 | 75.8 +/- 4.1 | 77.5 +/- 5.2 | 75.4 +/- 4.4 |
| Male | 224 (33.5) | 285 (42.7) | 332 (49.6) | 371 (55.4) | 1212 (45.3) |
| White | 526 (78.6) | 547 (81.9) | 554 (82.7) | 539 (80.4) | 2166 (80.9) |
| BMI, kg/m2 | 26.8 +/- 4.2 | 26.8 +/- 4.6 | 26.9 +/- 4.3 | 26.4 +/- 4.4 | 26.7 +/- 4.4 |
| Current smoker | 30 (4.5) | 53 (8) | 56 (8.4) | 81 (12.2) | 220 (8.3) |
| Heavy drinker | 44 (6.6) | 55 (8.2) | 47 (7) | 40 (6) | 186 (7) |
| Good/excellent health | 602 (90) | 585 (87.6) | 541 (80.7) | 487 (72.9) | 2215 (82.8) |
| Avg sitting systolic BP, mm Hg | 132.2 +/- 19.3 | 133.6 +/- 19.5 | 133.2 +/- 19.2 | 135.9 +/- 21.3 | 133.7 +/- 19.9 |
| EGFR | 80.0 +/- 14.4 | 73.7 +/- 14.3 | 68.1 +/- 15.5 | 55.7 +/- 17.0 | 69.4 +/- 17.7 |
| FEV1 | 2.2 +/- 0.6 | 2.1 +/- 0.7 | 2.1 +/- 0.7 | 2.0 +/- 0.7 | 2.1 +/- 0.7 |
| LDL (mg/dL) | 128.4 +/- 32.7 | 127.6 +/- 33.2 | 124.6 +/- 32.8 | 122.2 +/- 36.0 | 125.7 +/- 33.7 |
| HDL (mg/dL) | 56.4 +/- 15.9 | 53.6 +/- 14.5 | 52.6 +/- 15.5 | 50.4 +/- 15.1 | 53.3 +/- 15.4 |
| Triglyceride (mg/dL) | 140.3 +/- 71.3 | 136.2 +/- 77.0 | 142.3 +/- 84.8 | 154.3 +/- 96.3 | 143.2 +/- 83.1 |
| Teng 3MS score | 92.4 +/- 7.6 | 92.1 +/- 7.0 | 91.1 +/- 7.8 | 89.2 +/- 9.6 | 91.2 +/- 8.2 |
| Digit symbol substitution | 42.4 +/- 13.5 | 41.2 +/- 13.0 | 38.0 +/- 13.1 | 35.0 +/- 14.2 | 39.2 +/- 13.7 |
| 6m gait speed, m/s | 1.1 +/- 0.3 | 1.1 +/- 0.3 | 1.0 +/- 0.3 | 0.9 +/- 0.3 | 1.0 +/- 0.3 |
| Max grip strength, kg | 31.1 +/- 10.1 | 31.8 +/- 10.7 | 31.9 +/- 10.6 | 31.4 +/- 10.7 | 31.5 +/- 10.5 |
| 400m walk time | 314.2 +/- 63.7 | 315.4 +/- 74.7 | 316.7 +/- 69.2 | 328.9 +/- 84.4 | 318.0 +/- 72.5 |
| Six min walk distance | 1193.4 +/- 272.7 | 1201.3 +/- 286.4 | 1106.4 +/- 258.9 | 1055.1 +/- 296.3 | 1143.9 +/- 284.1 |
| Leg power | 91.7 +/- 32.3 | 98.6 +/- 38.6 | 94.0 +/- 31.3 | 93.0 +/- 31.5 | 94.3 +/- 33.8 |
| Log contrast sensitivity | 1.6 +/- 0.2 | 1.6 +/- 0.2 | 1.5 +/- 0.2 | 1.5 +/- 0.2 | 1.6 +/- 0.2 |
| Diabetes | 82 (12.6) | 83 (12.9) | 110 (17.2) | 157 (24.6) | 432 (16.8) |
| Anti-hypertensive medication | 276 (41.5) | 318 (47.7) | 365 (54.5) | 433 (64.8) | 1392 (52.1) |
| History of Coronary Heart Disease | 91 (13.6) | 123 (18.4) | 165 (24.6) | 212 (31.6) | 591 (22.1) |
| History of Atrial Fibrillation | 16 (2.4) | 38 (5.7) | 52 (7.8) | 77 (11.5) | 183 (6.8) |
| Two or more falls in follow-up | 38 (6.1) | 61 (9.6) | 57 (9.0) | 57 (9.7) | 213 (8.6) |

Note. Means +/- SD and n (%) presented.

### Cardiovascular disease

During mean follow-up of 21.9 years for the pooled cohort, there were 738 incident CHD events. For ASCVD, mean follow-up was 10.4 years with 670 incident events. Corresponding mean follow-up for HF was 10.7 years with 689 incident events. Of these, 235 were HFpEF and 231 were HFrEF, with the rest unclassified. Figure 1 shows the risk of cardiovascular outcomes by quartile of GDF-15. Under the fully adjusted model, the risk of CHD comparing quartile 4 to the referent quartile 1 was 1.47 (1.17, 1.83). The risk of atherosclerotic CVD was greater at the highest quartile of GDF-15 compared to the lowest quartile (HR: 1.56 [1.22, 1.98]). After adjusting for age, cohort, race, sex, BMI, smoking status, and heavy drinking, the risk of heart failure was nearly 2.5 times higher (HR: 2.54 [2.04, 3.17]) when comparing the highest quartile of GDF-15 to the lowest (eTable 1). After adding eGFR, FEV1, systolic blood pressure, and hypertensive medications, diabetes, prevalent CVD, prevalent stroke, prevalent claudication, prevalent atrial fibrillation, and CHD to the model, this was slightly attenuated but still associated with a higher risk of heart failure (HR: 2.09 [1.66, 2.64]). Each of these primary cardiovascular outcome associations were significant after Bonferroni correction. When comparing quartile 4 to quartile 1, the relationship between levels of GDF-15 and risk of HFpEF was significant after full adjustment (HR: 1.53 [1.04, 2.23]), but the association with the risk of HFrEF was not significant (HR: 1.21 [0.79, 1.84]) (eTable 3).

**Figure 1.**
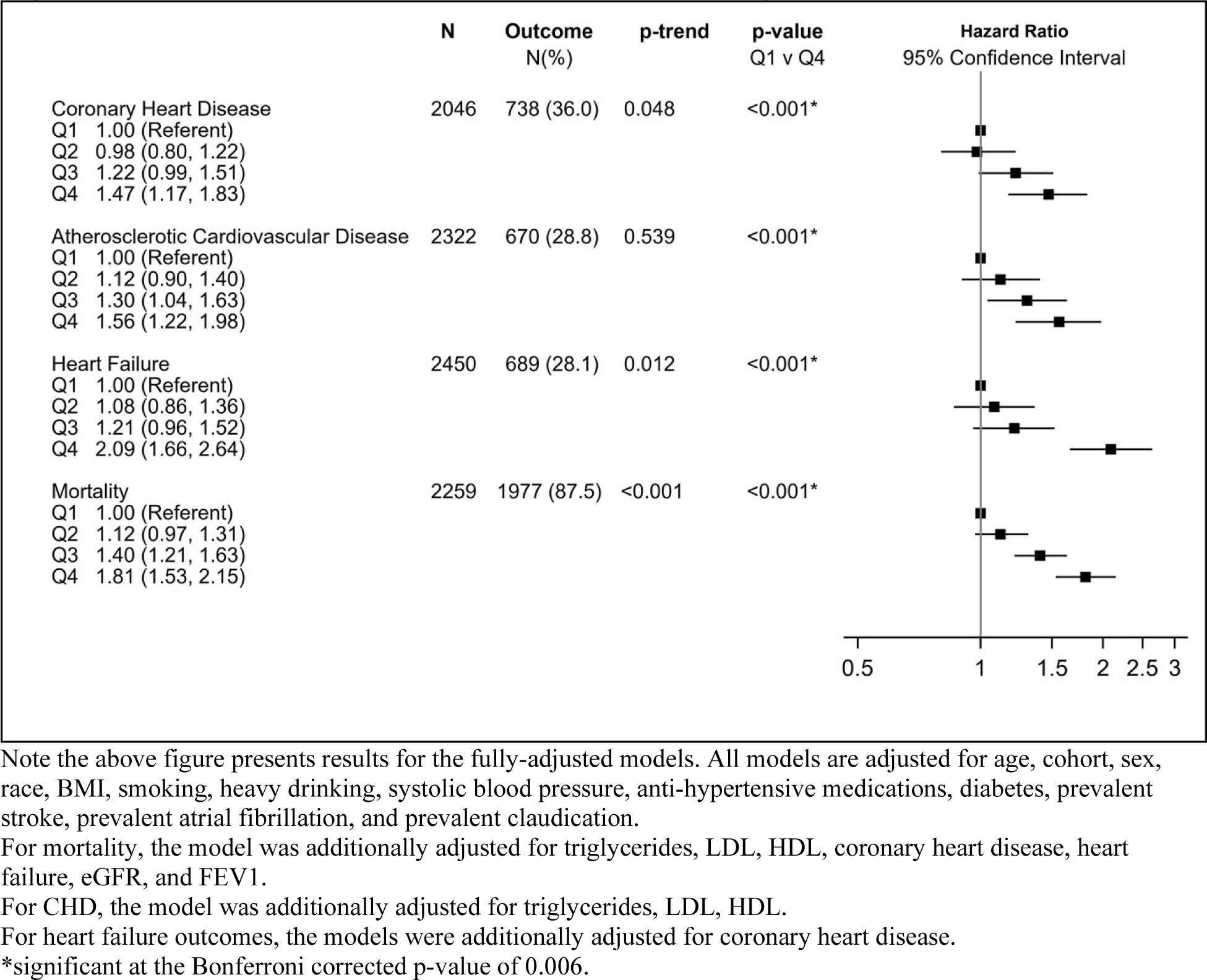
Risk for Cardiovascular Outcomes and Mortality

### Mortality

During the follow-up time of 11.5 years, there were 1,977 deaths in the pooled cohort. In the fully adjusted model, which is adjusted for age, cohort, sex, BMI, race, current smoker, heavy drinking, LDL, HDL, triglycerides, prevalent CHD, and prevalent stroke, systolic blood pressure, antihypertensive medications, diabetes, prevalent heart failure, claudication, FEV1, and eGFR those in the highest quartile of GDF 15 had a 1.8-fold greater risk of all-cause mortality than those in the lowest quartile (Figure 1).

### Dementia and Cognitive Performance

The risk of dementia was higher with increased GDF-15 levels (Figure 2). For the highest quartile compared to the lowest quartile of GDF-15, after adjustment for age, sex, race, BMI, smoking, and heavy drinking, the association was stronger in CHS (HR: 3.50 [1.97, 6.22]), in which dementia was adjudicated, compared to Health ABC (HR: 1.65 [1.13, 2.43]) in which the diagnosis of dementia was determined algorithmically. There was also a cross-sectional association between higher levels of GDF-15 and poor cognitive function measured by both Teng 3MS (p-trend = 0.015) and the DSS tests of cognitive performance (p-trend <0.001) (figure 3).

**Figure 2.**
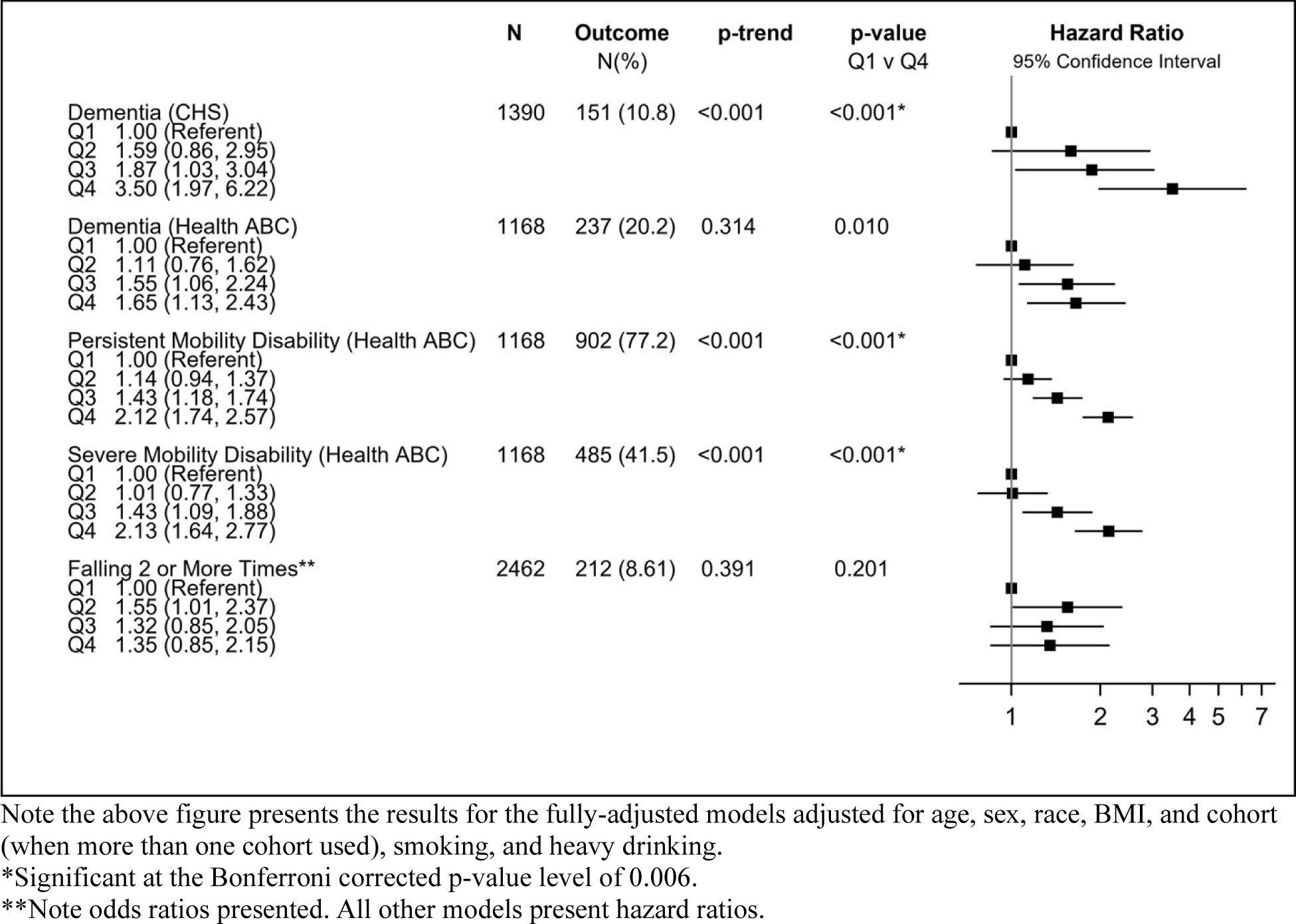
Risk or Odds for Cognitive and Mobility Disability Outcomes

**Figure 3.**
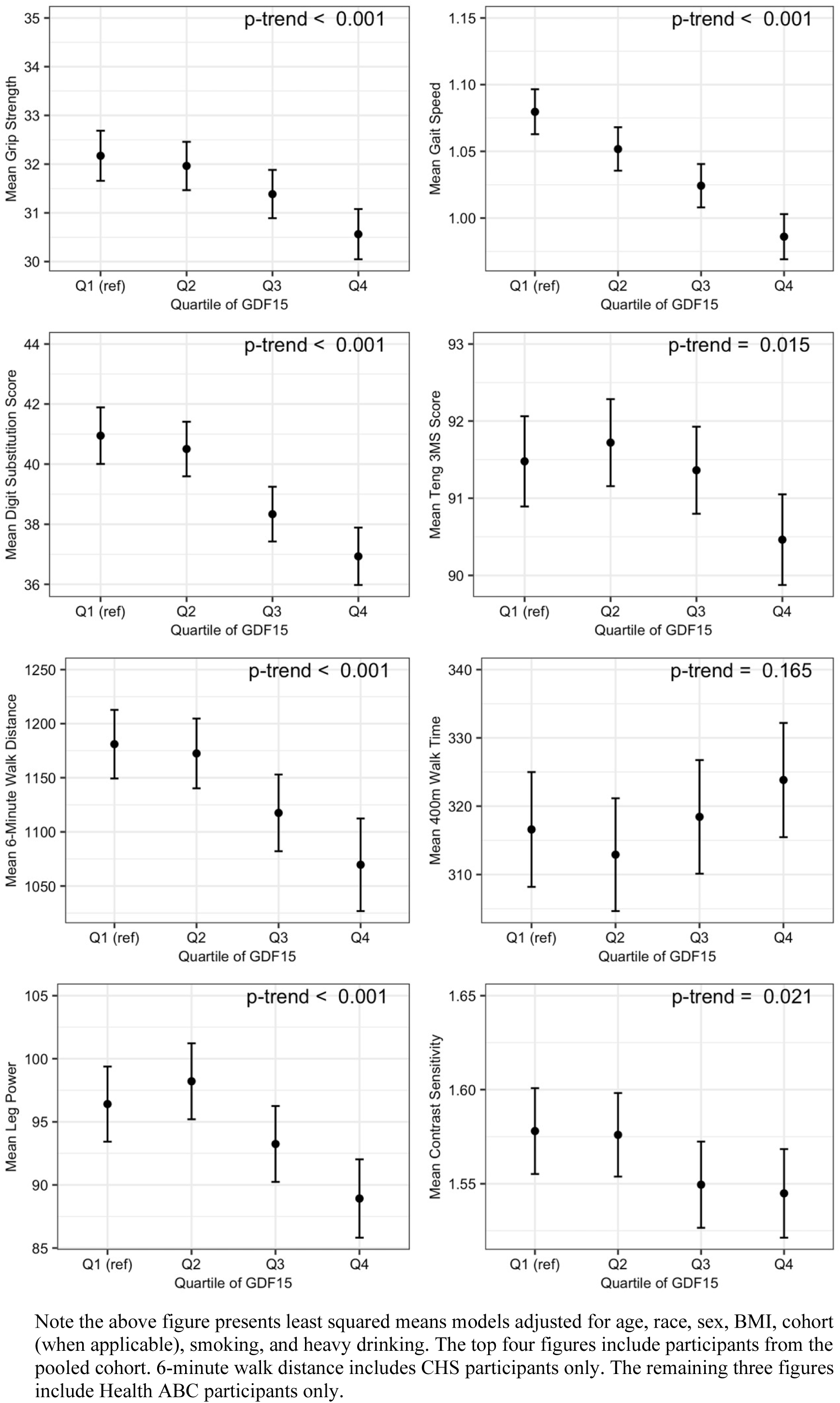
LS Means for Secondary Cognitive and Physical Performance Measures

### Mobility Disability

As shown in figure 2, higher levels of GDF-15 were associated with a greater risk of persistent mobility disability (HR: 2.12 [1.74, 2.57]) and severe mobility disability (HR: 2.13 [1.64, 2.77]).

### Falls and Physical Performance

Higher levels were not associated with an higher odds of multiple falls (figure 2). The cross-sectional association between GDF-15 and physical performance and strength was graded (figure 3); with increasing quartiles of GDF-15, we observed worse physical performance and strength (p for trend <0.001). Participants in the highest quartile of GDF-15 had lower knee extension strength; shorter six-minute walk distance; lower grip strength; and slower gait speed each with p for trend of <0.001 (figure 3). The 400m walk time was the only physical performance measure without a significant trend across quartiles of GDF-15.

### Other measurements

At higher levels of GDF-15 the contrast sensitivity score was lower with a p for trend of 0.02.

## Discussion

In our study of participants from the CHS and Health ABC cohorts, we found that GDF-15 was associated with mortality and a multitude of age-related outcomes and diseases, including impaired physical and cognitive performance, dementia, ASCVD, CHD, and HF. Our results are consistent with other studies that have assessed one clinical condition or certain aging-related tests of performance. In addition, we found novel associations between GDF-15 and persistent and severe mobility disability, as well as for secondary outcomes of vision loss, and leg power. Taken together, the diverse profile of effects indicates that GDF-15 is a biomarker of underlying aging processes that impact many tissues and physiologic processes.

Our results broadly support the work of other studies linking GDF-15 to worse physical performance. We observed that higher levels of GDF-15 were associated with worse physical performance outcomes for all measures except the 400m walk time and the odds of having two or more falls. We found a graded relationship between levels of GDF-15 and both grip and leg strength, consistent with a study of 1,096 community dwelling older adults that observed GDF-15 was cross-sectionally associated with weaker grip strength (6). We found that those with higher levels of GDF-15 had slower walking speeds as measured by the 6-minute walk test; however, there was no graded trend among levels of GDF-15 and 400m walk time. These 6-minute walk test results are consistent with a study of 194 participants from the Baltimore Longitudinal Study of Aging (17). The 400m walk results from that study are inconsistent with our finding, however. That study included participants aged 22 to 93; thus, the 400m walk results are not necessarily comparable to the older adults included in the present study. In another recent study that uses the data from CHS and the Framingham Offspring Study, gait speed using the 4-m walk (15-foot walk for CHS) was found to be associated with GDF-15 (19).

The association between GDF-1*5* levels and mobility limitations is consistent with other studies. A study of 660 older women and men used inability to complete the 400m walk to define risk of losing mobility and found that GDF-15 was significantly associated with the risk of losing mobility (20). Like that study, we found higher levels of GDF-15 were associated with higher risk of both persistent and severe mobility limitations as assessed by difficulty in walking ¼ mile or climbing 10 steps without resting. We did not, however, find an association between higher levels of GDF-15 and the odds of falling two or more times. To our knowledge this is the first study that has explored the association between GDF-15 and the odds of falling two or more times, and it is the first study to explore the association between GDF-15 and mobility limitations by our definitions.

These associations between GDF-15 and physical performance suggest a potential for GDF-15 as a neurotrophic factor for motor and sensory neurons. Consistent with this hypothesis, a study of GDF-15 knockout mice showed a progressive loss of motor axons and rotarod motor skills after 6 months (21). This supports the associations we observed between higher levels of GDF-15 and worse outcomes for physical performance.

GDF-15 may also present as a neurotrophic factor related to cognitive function. Consistent with prior findings, we observed that the risk of dementia was higher among those participants with high levels of GDF-15 (6, 22). Another study comparing Alzheimer dementia patients to healthy controls did not find a significant relationship between levels of GDF-15 and the risk of AD; however, that study did find a significant relationship when comparing Alzheimer dementia participants to offspring considered to be healthy (23). Beyond dementia, we found that DSST and Teng 3MS scores were worse at higher levels of GDF-15. DSST is a polyfactorial test that is highly sensitive to detecting cognitive impairment and requires both executive function and takes into consideration motor speed (24). This aligns with the physical performance findings above. At high levels of GDF-15, both physical and cognitive function are more impaired than they are at low levels again suggesting GDF-15’s role as a neurotrophic factor.

Our study results are consistent with prior prospective observational studies that have linked higher GDF-15 to increased risk of atherosclerotic CVD, HF, CHD and all-cause mortality (25,26,27). Although higher circulating GDF-15 level has been associated with increased incidence of stroke, no prior study has demonstrated this specifically for incident CHD events, as documented here. We also noted, consistent with a prior report, that higher GDF-15 was associated with risk of HFpEF, but not HFrEF (28).

Of note, a Mendelian randomization analysis did support a potentially causal association between genetically determined GDF-15 level and cardioembolic stroke (positive relationship), as well as CHD and MI (inverse relationship), though not HF or other stroke subtypes (29). It is known that GDF-15 levels increase in the setting of cellular stress and mitochondrial dysfunction, but that GDF-15 itself has healthful anti-oxidant, anti-inflammatory and anti-apoptotic properties (30). The basis for documented differences in direction of genetic associations is unclear, but may reflect the balance of salutary and as yet undefined deleterious actions of the protein or associated pathways. Regardless, the present findings extend knowledge of the role of GDF-15 as a marker of adverse risk for CHD in addition to HF and atherosclerotic CVD more broadly, the incremental predictive value of which will require formal evaluation in studies of risk prediction. Further elucidation of GDF-15’s biological actions and therapeutic potential will require additional study through MR approaches, laboratory work, and ultimately randomized trials.

In the present study, we also observed associations between GDF-15 and other conditions. In our exploration of these associations, depression was the only outcome without a significant trend across levels of GDF-15. As a summary measure, higher levels of GDF-15 were associated with lower odds of self-reporting good or excellent health.

Despite the breadth of associations found, the precise mechanisms of GDF-15 effects are unknown. In addition, the origin of circulating GDF-15 in older adults is not certain. Two studies using unbiased proteomics to identify biomarkers of cell senescence using in vitro models of senescence and clinical conditions, such as Schafer et al. found that GDF-15 was the only protein with significantly increased abundance in all models of senescence (4). If an increased level GDF-15 is a marker of cell senescence, this could account for its pleiotropic associations with many aging-related diseases and mortality. That might also account for the numerous associations that we have found in cohorts of older adults. This raises the possibility that GDF-15 levels could be a useful marker for assessment of senolytic treatments, a possibility that will require further investigation in randomized trials.

Our study has several strengths. The cohorts have multiple standardized assessments of physical and cognitive performance along with adjudication of disease endpoints. The populations are racially diverse with equal numbers of men and women. To our knowledge, it is the most comprehensive examination of the range of diverse aging-related endpoints in older adults. There are also several limitations, including measurement error and selection bias. For certain outcomes, data was only available from one cohort. Our findings may not be generalizable to all older adults or to racially/ethnically distinct or younger populations.

In conclusion, we identified novel associations of GDF-15 with incident mobility limitation and coronary heart disease. Our findings lend further support to the concept that increased GDF-15 levels are associated with an increased risk of multiple aging-related phenotypes and outcomes that highlighting its status as a marker of underlying processes of biological aging. Additional study is needed to determine whether GDF-15 levels are useful in risk prediction for the new associated outcomes we have documented. GDF-15 could serve as a useful aging biomarker for evaluation of senolytic therapies. Interventions on the pathways that lower GDF-15 levels in older adults could provide new approaches to extending healthy life expectancy.

## Data Availability

All data produced in the present study are available upon reasonable request to authors.

## Acknowledgements

CHS was supported by R01 AG053325 from the National Institute on Aging; and by contracts HHSN268201200036C, HHSN268200800007C, HHSN268201800001C, N01HC55222, N01HC85079, N01HC85080, N01HC85081, N01HC85082, N01HC85083, N01HC85086, 75N92021D00006, and grants U01HL080295 and U01HL130114 from the National Heart, Lung, and Blood Institute (NHLBI), with additional contribution from the National Institute of Neurological Disorders and Stroke (NINDS). Additional support was provided by R01AG023629 from the National Institute on Aging (NIA). A full list of principal CHS investigators and institutions can be found at CHS-NHLBI.org. Health ABC was supported by National Institute on Aging (NIA) Contracts N01-AG-6-2101; N01-AG-6-2103; N01-AG-6-2106; NIA grant R01-AG028050, and NINR grant R01-NR012459. The research was funded in part by the Intramural Research Program of the NIH, National Institute on Aging.

---

**eTable 1.**
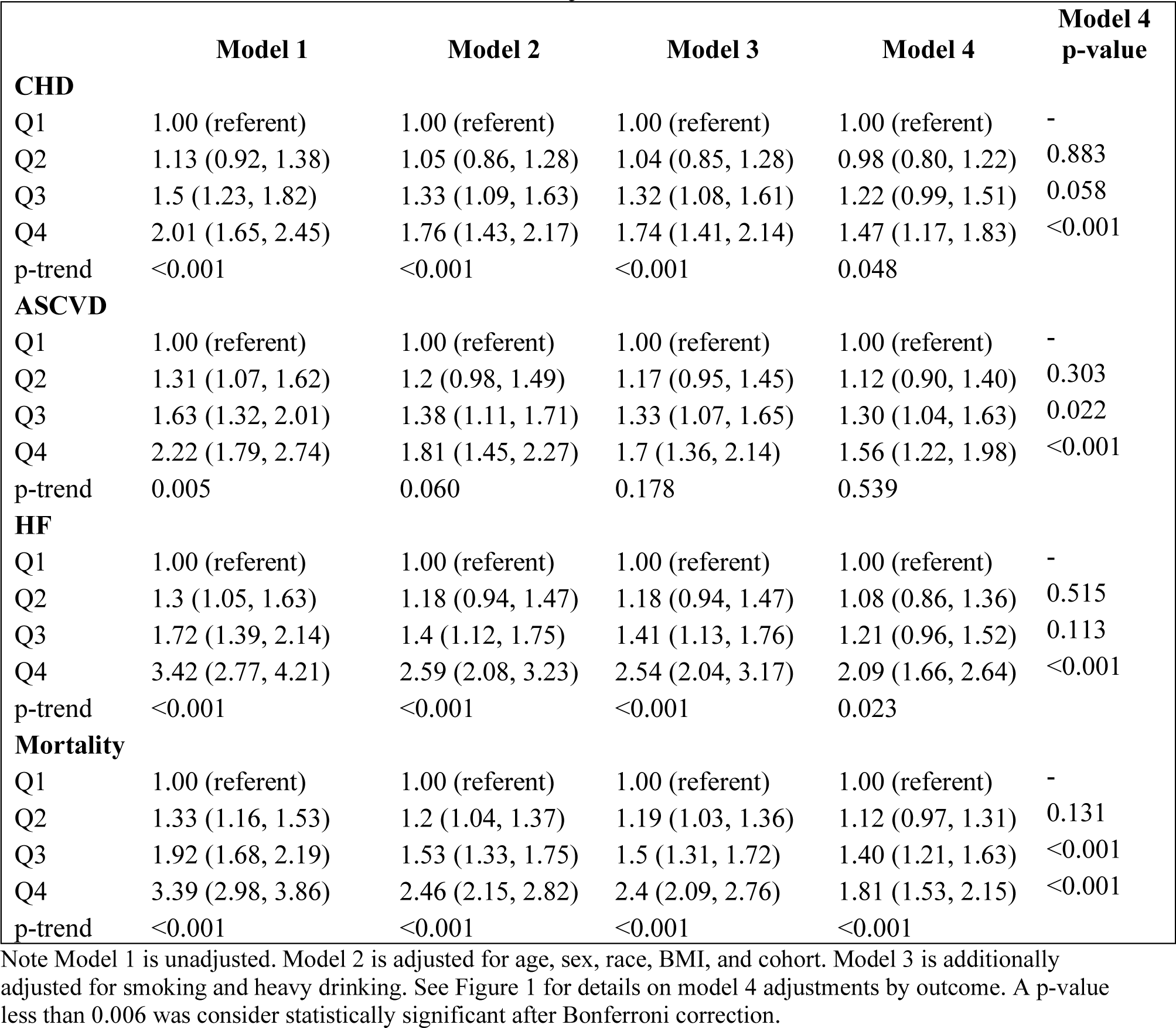
Associations (hazard ratios and 95% confidence intervals) for mortality and cardiovascular outcomes with and without adjustment.

**eTable 2.**
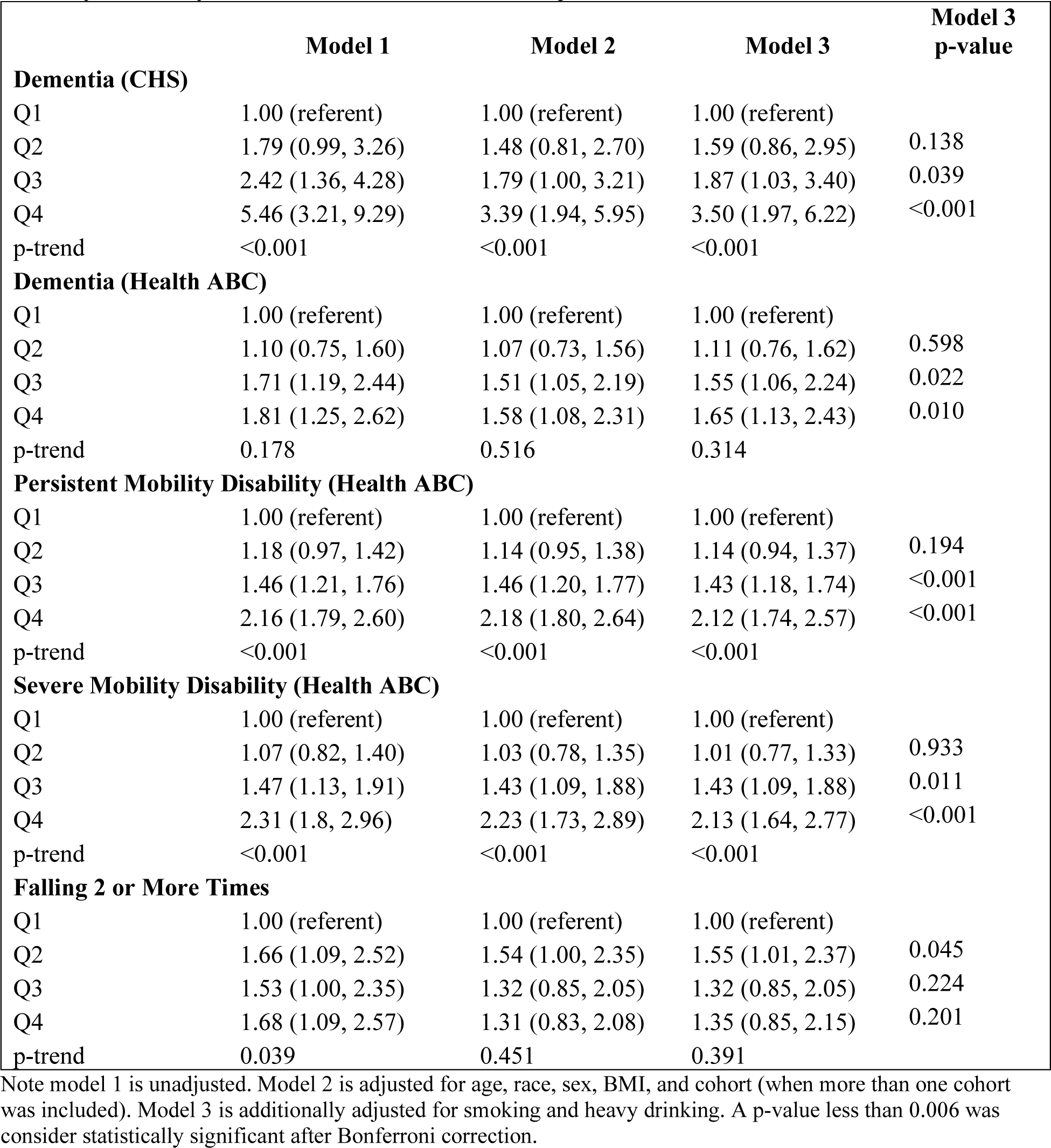
Associations (hazard or odds ratios and 95% confidence intervals) for dementia, mobility disability, and falls with and without adjustment.

**eTable 3.**
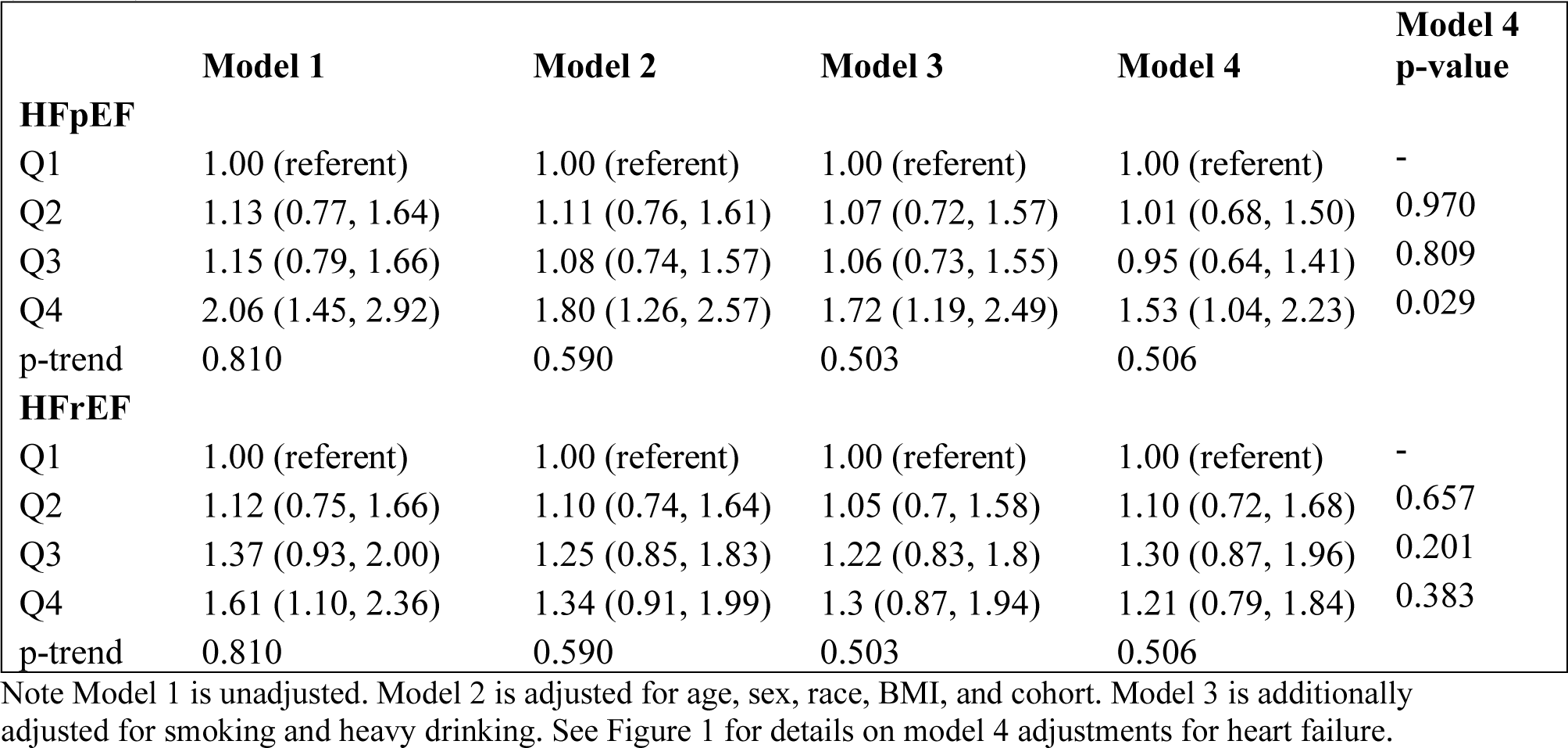
Associations (hazard ratios and 95% confidence intervals) for heart failure with preserved ejection fraction (HFpEF) and heart failure with reduced ejection fraction (HFrEF)

