## Supplemental tables 1-3 for "Associations of Serum GDF-15 Levels with Physical Performance, Mobility Disability, Cognition, Cardiovascular Disease, and Mortality in Older Adults"

**eTable 1. Associations (hazard ratios and 95% confidence intervals) for mortality and cardiovascular outcomes with and without adjustment**

| | Model 1 | Model 2 | Model 3 | Model 4 | Model 4 p-value |
|---|---|---|---|---|---|
| **CHD** | | | | | |
| Q1 | 1.00 (referent) | 1.00 (referent) | 1.00 (referent) | 1.00 (referent) | - |
| Q2 | 1.13 (0.92, 1.38) | 1.05 (0.86, 1.28) | 1.04 (0.85, 1.28) | 0.98 (0.80, 1.22) | 0.883 |
| Q3 | 1.5 (1.23, 1.82) | 1.33 (1.09, 1.63) | 1.32 (1.08, 1.61) | 1.22 (0.99, 1.51) | 0.058 |
| Q4 | 2.01 (1.65, 2.45) | 1.76 (1.43, 2.17) | 1.74 (1.41, 2.14) | 1.47 (1.17, 1.83) | <0.001 |
| p-trend | <0.001 | <0.001 | <0.001 | 0.048 | |
| **ASCVD** | | | | | |
| Q1 | 1.00 (referent) | 1.00 (referent) | 1.00 (referent) | 1.00 (referent) | - |
| Q2 | 1.31 (1.07, 1.62) | 1.2 (0.98, 1.49) | 1.17 (0.95, 1.45) | 1.12 (0.90, 1.40) | 0.303 |
| Q3 | 1.63 (1.32, 2.01) | 1.38 (1.11, 1.71) | 1.33 (1.07, 1.65) | 1.30 (1.04, 1.63) | 0.022 |
| Q4 | 2.22 (1.79, 2.74) | 1.81 (1.45, 2.27) | 1.7 (1.36, 2.14) | 1.56 (1.22, 1.98) | <0.001 |
| p-trend | 0.005 | 0.060 | 0.178 | 0.539 | |
| **HF** | | | | | |
| Q1 | 1.00 (referent) | 1.00 (referent) | 1.00 (referent) | 1.00 (referent) | - |
| Q2 | 1.3 (1.05, 1.63) | 1.18 (0.94, 1.47) | 1.18 (0.94, 1.47) | 1.08 (0.86, 1.36) | 0.515 |
| Q3 | 1.72 (1.39, 2.14) | 1.4 (1.12, 1.75) | 1.41 (1.13, 1.76) | 1.21 (0.96, 1.52) | 0.113 |
| Q4 | 3.42 (2.77, 4.21) | 2.59 (2.08, 3.23) | 2.54 (2.04, 3.17) | 2.09 (1.66, 2.64) | <0.001 |
| p-trend | <0.001 | <0.001 | <0.001 | 0.023 | |
| **Mortality** | | | | | |
| Q1 | 1.00 (referent) | 1.00 (referent) | 1.00 (referent) | 1.00 (referent) | - |
| Q2 | 1.33 (1.16, 1.53) | 1.2 (1.04, 1.37) | 1.19 (1.03, 1.36) | 1.12 (0.97, 1.31) | 0.131 |
| Q3 | 1.92 (1.68, 2.19) | 1.53 (1.33, 1.75) | 1.5 (1.31, 1.72) | 1.40 (1.21, 1.63) | <0.001 |
| Q4 | 3.39 (2.98, 3.86) | 2.46 (2.15, 2.82) | 2.4 (2.09, 2.76) | 1.81 (1.53, 2.15) | <0.001 |
| p-trend | <0.001 | <0.001 | <0.001 | <0.001 | |

Note Model 1 is unadjusted. Model 2 is adjusted for age, sex, race, BMI, and cohort. Model 3 is additionally adjusted for smoking and heavy drinking. See Figure 1 for details on model 4 adjustments by outcome. A p-value less than 0.006 was consider statistically significant after Bonferroni correction.

**eTable 2. Associations (hazard or odds ratios and 95% confidence intervals) for dementia, mobility disability, and falls with and without adjustment**

| | Model 1 | Model 2 | Model 3 | Model 3 p-value |
|---|---|---|---|---|
| **Dementia (CHS)** | | | | |
| Q1 | 1.00 (referent) | 1.00 (referent) | 1.00 (referent) | |
| Q2 | 1.79 (0.99, 3.26) | 1.48 (0.81, 2.70) | 1.59 (0.86, 2.95) | 0.138 |
| Q3 | 2.42 (1.36, 4.28) | 1.79 (1.00, 3.21) | 1.87 (1.03, 3.40) | 0.039 |
| Q4 | 5.46 (3.21, 9.29) | 3.39 (1.94, 5.95) | 3.50 (1.97, 6.22) | <0.001 |
| p-trend | <0.001 | <0.001 | <0.001 | |
| **Dementia (Health ABC)** | | | | |
| Q1 | 1.00 (referent) | 1.00 (referent) | 1.00 (referent) | |
| Q2 | 1.10 (0.75, 1.60) | 1.07 (0.73, 1.56) | 1.11 (0.76, 1.62) | 0.598 |
| Q3 | 1.71 (1.19, 2.44) | 1.51 (1.05, 2.19) | 1.55 (1.06, 2.24) | 0.022 |
| Q4 | 1.81 (1.25, 2.62) | 1.58 (1.08, 2.31) | 1.65 (1.13, 2.43) | 0.010 |
| p-trend | 0.178 | 0.516 | 0.314 | |
| **Persistent Mobility Disability (Health ABC)** | | | | |
| Q1 | 1.00 (referent) | 1.00 (referent) | 1.00 (referent) | |
| Q2 | 1.18 (0.97, 1.42) | 1.14 (0.95, 1.38) | 1.14 (0.94, 1.37) | 0.194 |
| Q3 | 1.46 (1.21, 1.76) | 1.46 (1.20, 1.77) | 1.43 (1.18, 1.74) | <0.001 |
| Q4 | 2.16 (1.79, 2.60) | 2.18 (1.80, 2.64) | 2.12 (1.74, 2.57) | <0.001 |
| p-trend | <0.001 | <0.001 | <0.001 | |
| **Severe Mobility Disability (Health ABC)** | | | | |
| Q1 | 1.00 (referent) | 1.00 (referent) | 1.00 (referent) | |
| Q2 | 1.07 (0.82, 1.40) | 1.03 (0.78, 1.35) | 1.01 (0.77, 1.33) | 0.933 |
| Q3 | 1.47 (1.13, 1.91) | 1.43 (1.09, 1.88) | 1.43 (1.09, 1.88) | 0.011 |
| Q4 | 2.31 (1.8, 2.96) | 2.23 (1.73, 2.89) | 2.13 (1.64, 2.77) | <0.001 |
| p-trend | <0.001 | <0.001 | <0.001 | |
| **Falling 2 or More Times** | | | | |
| Q1 | 1.00 (referent) | 1.00 (referent) | 1.00 (referent) | |
| Q2 | 1.66 (1.09, 2.52) | 1.54 (1.00, 2.35) | 1.55 (1.01, 2.37) | 0.045 |
| Q3 | 1.53 (1.00, 2.35) | 1.32 (0.85, 2.05) | 1.32 (0.85, 2.05) | 0.224 |
| Q4 | 1.68 (1.09, 2.57) | 1.31 (0.83, 2.08) | 1.35 (0.85, 2.15) | 0.201 |
| p-trend | 0.039 | 0.451 | 0.391 | |

Note model 1 is unadjusted. Model 2 is adjusted for age, race, sex, BMI, and cohort (when more than one cohort was included). Model 3 is additionally adjusted for smoking and heavy drinking. A p-value less than 0.006 was consider statistically significant after Bonferroni correction.

**eTable 3. Associations (hazard ratios and 95% confidence intervals) for heart failure with preserved ejection fraction (HFpEF) and heart failure with reduced ejection fraction (HFrEF)**

| | **Model 1** | **Model 2** | **Model 3** | **Model 4** | **Model 4 p-value** |
|---|---|---|---|---|---|
| **HFpEF** | | | | | |
| Q1 | 1.00 (referent) | 1.00 (referent) | 1.00 (referent) | 1.00 (referent) | - |
| Q2 | 1.13 (0.77, 1.64) | 1.11 (0.76, 1.61) | 1.07 (0.72, 1.57) | 1.01 (0.68, 1.50) | 0.970 |
| Q3 | 1.15 (0.79, 1.66) | 1.08 (0.74, 1.57) | 1.06 (0.73, 1.55) | 0.95 (0.64, 1.41) | 0.809 |
| Q4 | 2.06 (1.45, 2.92) | 1.80 (1.26, 2.57) | 1.72 (1.19, 2.49) | 1.53 (1.04, 2.23) | 0.029 |
| p-trend | 0.810 | 0.590 | 0.503 | 0.506 | |
| **HFrEF** | | | | | |
| Q1 | 1.00 (referent) | 1.00 (referent) | 1.00 (referent) | 1.00 (referent) | - |
| Q2 | 1.12 (0.75, 1.66) | 1.10 (0.74, 1.64) | 1.05 (0.7, 1.58) | 1.10 (0.72, 1.68) | 0.657 |
| Q3 | 1.37 (0.93, 2.00) | 1.25 (0.85, 1.83) | 1.22 (0.83, 1.8) | 1.30 (0.87, 1.96) | 0.201 |
| Q4 | 1.61 (1.10, 2.36) | 1.34 (0.91, 1.99) | 1.3 (0.87, 1.94) | 1.21 (0.79, 1.84) | 0.383 |
| p-trend | 0.810 | 0.590 | 0.503 | 0.506 | |

Note Model 1 is unadjusted. Model 2 is adjusted for age, sex, race, BMI, and cohort. Model 3 is additionally adjusted for smoking and heavy drinking. See Figure 1 for details on model 4 adjustments for heart failure.
